# Application and validation of an algorithmic classification of early impairment in cognitive performance

**DOI:** 10.1101/2023.02.04.23285477

**Authors:** Yurun Cai, Jennifer A. Schrack, Yuri Agrawal, Nicole M. Armstrong, Amal Wanigatunga, Melissa Kitner-Triolo, Abhay Moghekar, Luigi Ferrucci, Eleanor M. Simonsick, Susan M. Resnick, Alden L. Gross

**Affiliations:** Department of Epidemiology, Johns Hopkins Bloomberg School of Public Health, Baltimore, MD, USA; Department of Health and Community Systems, University of Pittsburgh School of Nursing, Pittsburgh, PA, USA; Center on Aging and Health, Johns Hopkins School of Medicine, Baltimore, MD, USA; Department of Otolaryngology - Head and Neck Surgery, Johns Hopkins School of Medicine, Baltimore, MD, USA; Department of Psychiatry and Human Behavior, Warren Alpert Medical School, Brown University, Providence, RI, USA; Intramural Research Program, National Institute on Aging, Baltimore, MD, USA; Department of Neurology, Johns Hopkins School of Medicine, Baltimore, MD, USA

**Author notes:** **Corresponding author:** Yurun Cai, PhD, 615 N. Wolfe St., Baltimore, MD 21205, **Present address:** 415 Victoria Building, 3500 Victoria St, Pittsburgh, PA 15261.

**Keywords:** Alzheimer’s Disease, Cognitive Dysfunction, Neuropsychological Tests, Classification, Validation Study, Longitudinal Studies

## Abstract

**Objective:** Due to the long prodromal period for dementia pathology, approaches are needed to detect cases before clinically recognizable symptoms are apparent, by which time it is likely too late to intervene. This study contrasted two theoretically-based algorithms for classifying early cognitive impairment (ECI) in adults aged ≥50 enrolled in the Baltimore Longitudinal Study of Aging.

**Method:** Two ECI algorithms were defined as poor performance (1 standard deviation [SD] below age-, sex-, race-, and education-specific means) in: (1) Card Rotations or California Verbal Learning Test (CVLT) immediate recall and (2) ≥1 (out of 2) memory or ≥3 (out of 6) non-memory tests. We evaluated concurrent criterion validity against consensus diagnoses of mild cognitive impairment (MCI) or dementia and global cognitive scores using receiver operating characteristic (ROC) curve analysis. Predictive criterion validity was evaluated using Cox proportional hazards models to examine the associations between algorithmic status and future adjudicated MCI/dementia.

**Results:** Among 1,851 participants (mean age=65.2±11.8 years, 50% women, 74% white), the two ECI algorithms yielded comparably moderate concurrent criterion validity with adjudicated MCI/dementia. For predictive criterion validity, the algorithm based on impairment in Card Rotations or CVLT immediate recall was the better predictor of MCI/dementia (HR=3.53, 95%CI: 1.59-7.84) over 12.3 follow-up years.

**Conclusions:** Impairment in visuospatial ability or memory may be capable of detecting early cognitive changes in the preclinical phase among cognitively normal individuals.

## Introduction

Alzheimer’s disease (AD) is a neurodegenerative disease that interferes with daily activities in its later stages. Identification of early progression is critical to identify risk factors and to properly evaluate interventions to delay clinical onset. Progression to AD is hypothesized to begin with a preclinical phase characterized by normal cognitive ability, up to 20 years prior to a dementia diagnosis. ^1–3^ Thus, accurate and stable diagnostic criteria for identifying early cognitive changes prior to clinically recognizable symptoms of dementia are crucial for purposes of targeting preventive interventions most likely to slow pathological progression.

A wide range of neuropsychological measures have been leveraged in epidemiologic studies for the classification of dementia and mild cognitive impairment (MCI). There is a plethora of different algorithms developed based on poor performance in cognitive and everyday functional measures to classify MCI or dementia in lieu of clinical judgement, frequently motivated by various psychiatric ^2,3^ or neuropsychological traditions ^4–6^. As episodic memory impairment is most seen in MCI patients who progress to AD, conventional Petersen criteria defined MCI is based on performance >1.5 standard deviation (SD) below age-appropriate norms on a single memory test ^7^. This approach has been expanded to multiple other cognitive domains (e.g., executive, language) and requires multiple tests, within each cognitive domain >1 SD below age-appropriate norms to balance sensitivity and specificity. ^4^ Although neuropsychological criteria have been validated in multiple cohort studies for MCI classification, ^5,6^ few studies have carefully considered which cognitive tests should be included to detect early cognitive decline in preclinical stages of dementia before the symptomatic phase. ^8^ Specifically, visuospatial function has not been considered as a separate domain in these algorithms, however, a recent study reported that visuospatial ability measured using the Card Rotations test showed the earliest changes in rate of decline at 15.5 years before AD diagnosis, followed by episodic memory where changes were detected up to 11.7 years before AD diagnosis^9^. These findings make some biological sense, given that visuospatial ability as measured by Card Rotations are thought to be controlled in the brain’s precuneous and retrosplenial cortex, atrophy in which is an early risk factor for ADRD.^9–11^ Thus, incorporating visual function tests in algorithms may help identify individuals who are at high risk of developing ADRD in early stages. ^12^

We leveraged data from the Baltimore Longitudinal Study of Aging (BLSA), which has followed people for up to 33 years prior to dementia diagnosis starting as early as 1986. In the present study, we aim to contrast two psychometrically defined algorithms for classifying early cognitive impairment (ECI) in middle-aged and older adults enrolled in the BLSA. One algorithm was developed based on visual spatial and episodic memory which have showed early decline in progression to ADRD.^9^ Another algorithm used conventional neuropsychological criteria to detect early amnestic or nonamnestic cognitive decline which we hypothesize may predict all-cause dementia. By comparing different algorithmic classification criteria, we aim to identify optimal classification criteria for early identification of older adults with risk of MCI and dementia. We evaluated these algorithms by comparing concurrent and predictive criterion validation against consensus diagnoses and global measures of cognitive and functional impairment.

## Methods

The BLSA is a longitudinal cohort study established in 1958 and conducted by the National Institute on Aging Intramural Research Program. The study aims to explore the interdependence of aging and disease processes and their mutual impact on physical and cognitive function. A detailed description of the study design is available ^15^. The study continuously recruits community-dwelling volunteers free of major chronic conditions and cognitive and functional impairment at the time of enrollment. Participants are followed for health characteristics, cognitive assessments, and physical function testing every 1-4 years depending on age (every 4 years for age <60, every 2 years for age 60-79, and annually for age ≥80). The present study includes 1,880 participants aged ≥50 years who underwent cognitive testing from 1993 through 2019. Informed consent was obtained from all participants. The study protocol was approved by the National Institutes of Health Intramural Institutional Review Board.

### Neuropsychological tests

A wide variety of cognitive tests are administered in the BLSA. In the current study, attention and executive function were assessed using the Digit Span Forward and Backward subtests in the Wechsler Adult Intelligence Scale – Revised (WAIS-R) ^16^. Visual memory was measured using the Benton Visual Retention Test (BVRT) ^17^. Language was assessed using the 60-item Boston Naming Test (BNT-60) ^18^ and Similarities from the Wechsler Adult Intelligence Scale (WAIS) ^16^. Visuospatial ability was measured using the difference between the number of correctly and incorrectly classified objects on a modified version of the Card Rotations test developed by the Educational Testing Service ^19^. Verbal episodic memory was measured using the immediate (total number of items recalled across five trials) and long-delay free recall in the California Verbal Learning Test (CVLT) ^20^.

### Algorithmic classification of ECI

The ECI algorithms were determined based on the Preclinical AD Consortium but tailored for the BLSA sample depending on the number of neuropsychological tests administered in each cognitive domain and cutoff points used ^8^. Briefly, poor cognitive performance was operationalized as 1 SD below age-, sex-, race- (white vs nonwhite), and education-specific means. The race adjustment in addition to education is based on the consideration that education may not indicate the same level of educational attainment or intellectual exposure for different racial groups in the US particularly for those growing up in the 60’s and 70’s. According to previous literature using Jak/Bondi comprehensive criteria, ECI classification was determined based on memory and non-memory domains ^5,6^. In this study, we classified CVLT immediate and long-delay free recall as memory tests and other neuropsychological tests as non-memory tests. As previous findings suggesting that visuospatial ability measured by Card Rotations test and CVLT immediate recall showed the earliest changes in cognitive decline during preclinical stage of AD ^9,21^, we explored the algorithms using visuospatial ability and immediate recall for ECI classification. Thus, in this study, two ECI classification algorithms were developed and compared: (1) poor performance in Card Rotations or CVLT immediate recall and (2) poor performance in ≥1 (out of 2) memory or ≥3 (out of 6) non-memory tests.

### Global cognitive and functional scores

Three measures of global cognitive and/or functional status were used. Global mental status was assessed using the Mini-Mental State Examination (MMSE) ^22^. The Blessed Information Memory Concentration (BIMC) test is a mental status instrument that has been widely used in clinical populations and research studies ^23^. CDR Sum of Boxes (CDR-SB) is a global cognitive and functional assessment of six domains: memory, orientation, judgement/problem solving, community affairs, home/hobbies, and personal care ^24,25^.

### Adjudicated diagnosis of MCI/dementia

Participants with BIMC test score ≥4 or CDR score ≥0.5 were reviewed at consensus diagnostic conferences. Experienced clinicians diagnosed MCI based on Petersen criteria ^7^ and dementia based on Diagnostic and Statistical Manual of Mental Disorders, revised third edition criteria ^26,27^.

### Covariates

Sociodemographic characteristics including age, sex, race, and years of education were collected from a health interview. Race was categorized into white and non-white (e.g., Black, American Indian/Alaska Native, Asian/Pacific Islanders).

### Statistical Analysis

Sample characteristics including baseline age, sex, race, and years of education were summarized into frequencies and percentages or means and standard deviations.

First, we evaluated concurrent validity of the algorithms with concurrent consensus diagnoses of MCI/dementia **(Table 1)**. The receiver operating characteristic (ROC) curve analysis was used to calculate area under the curve (AUC), sensitivity, and specificity. All BLSA visits among all eligible participants with available cognitive data were included in the analysis. We also examined whether each ECI algorithmic classification was concurrently correlated with MMSE score and other global cognitive and functioning scores (BIMC and CDR-SB scores). Dichotomous global scores were used for the analysis, with cutoff points determined as MMSE ≤26 ^28^, BIMC ≥4 ^29^, and CDR-SB ≥0.5 ^24^.

**Table 1.** Concurrent criterion validity for each algorithmic classification of early cognitive impairment (ECI) with consensus diagnosis and global cognitive scores.

| **Algorithmic classification of early cognitive impairment** | **AUC** | **Sensitivity** | **Specificity** | **Kappa** | **N\*** | **TP** | **FP** | **FN** | **TN** |
| --- | --- | --- | --- | --- | --- | --- | --- | --- | --- |
| **Consensus diagnosis of MCI/dementia** |  |  |  |  |  |  |  |  |  |
| impairment in Card Rotation or CVLT immediate recall | 0.631 | 0.52 | 0.74 | 0.03 | 9466 | 67 | 2443 | 61 | 6895 |
| impairment in 1+ memory or 3+ non-memory tests | 0.703 | 0.65 | 0.75 | 0.04 | 9524 | 85 | 2327 | 45 | 7067 |
| **Global cognitive scores** |  |  |  |  |  |  |  |  |  |
| **MMSE score ($\leq 26$)** |  |  |  |  |  |  |  |  |  |
| impairment in Card Rotation or CVLT immediate recall | 0.640 | 0.51 | 0.77 | 0.17 | 7551 | 416 | 1563 | 398 | 5174 |
| impairment in 1+ memory or 3+ non-memory tests | 0.670 | 0.56 | 0.78 | 0.21 | 7551 | 457 | 1492 | 357 | 5245 |
| **CDR-SB score ($\geq 0.5$)** |  |  |  |  |  |  |  |  |  |
| impairment in Card Rotation or CVLT immediate recall | 0.586 | 0.36 | 0.81 | 0.18 | 2073 | 264 | 254 | 466 | 1089 |
| impairment in 1+ memory or 3+ non-memory tests | 0.610 | 0.40 | 0.82 | 0.23 | 2073 | 295 | 248 | 435 | 1095 |
| **BIMC score ($\geq 4$)** |  |  |  |  |  |  |  |  |  |
| impairment in Card Rotation or CVLT immediate recall | 0.623 | 0.48 | 0.76 | 0.16 | 8892 | 506 | 1860 | 543 | 5983 |
| impairment in 1+ memory or 3+ non-memory tests | 0.659 | 0.54 | 0.78 | 0.21 | 8892 | 565 | 1727 | 484 | 6116 |

*Note.* MCI=mild cognitive impairment. CVLT=California Verbal Learning Test. MMSE=Mini-Mental State Examination. CDR-SB=Clinical Dementia Rating - Sum of Boxes. BIMC=Blessed Information Memory Concentration test. AUC=area under the curve. TP=true positive. FP=false positive. FN=false negative. TN=true negative.
Poor performance in global cognitive score was determined based on $MMSE \leq 26$ , $BIMC \geq 4$ , and $CDR-SB \geq 0.5$ .
\*Number of visits among all eligible participants with available cognitive data.

Second, we evaluated predictive criterion validity of each algorithm with future progression to adjudicated MCI/dementia **(Table 2)**. Cox proportional hazards models were used to model associations between the earliest ECI status determined based on each algorithmic classification and time to adjudicated MCI/dementia during follow up. Participants who had MCI/dementia diagnosis at the first visit with available cognitive data were excluded from this analysis.

**Table 2.**
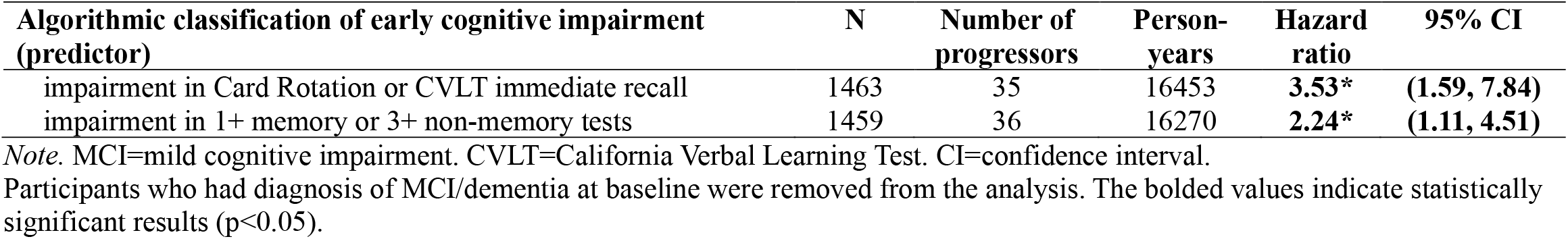
Predictive criterion validity for each algorithmic classification of early cognitive impairment (ECI) at baseline predicting progression to consensus diagnosis of MCI/dementia during follow up.

| **Algorithmic classification of early cognitive impairment (predictor)** | **N** | **Number of progressors** | **Person-years** | **Hazard ratio** | **95% CI** |
| --- | --- | --- | --- | --- | --- |
| impairment in Card Rotation or CVLT immediate recall | 1463 | 35 | 16453 | **3.53\*** | **(1.59, 7.84)** |
| impairment in 1+ memory or 3+ non-memory tests | 1459 | 36 | 16270 | **2.24\*** | **(1.11, 4.51)** |

*Note.* MCI=mild cognitive impairment. CVLT=California Verbal Learning Test. CI=confidence interval.
Participants who had diagnosis of MCI/dementia at baseline were removed from the analysis. The bolded values indicate statistically significant results ( $p < 0.05$ ).

Third, we additionally examined the association between baseline MMSE, BIMC, and CDR-SB scores and time to first algorithmically defined ECI during follow up, adjusted for age, sex, race, and years of education **(Table 3)**. Participants who already had ECI at baseline were excluded from this analysis.

**Table 3.**
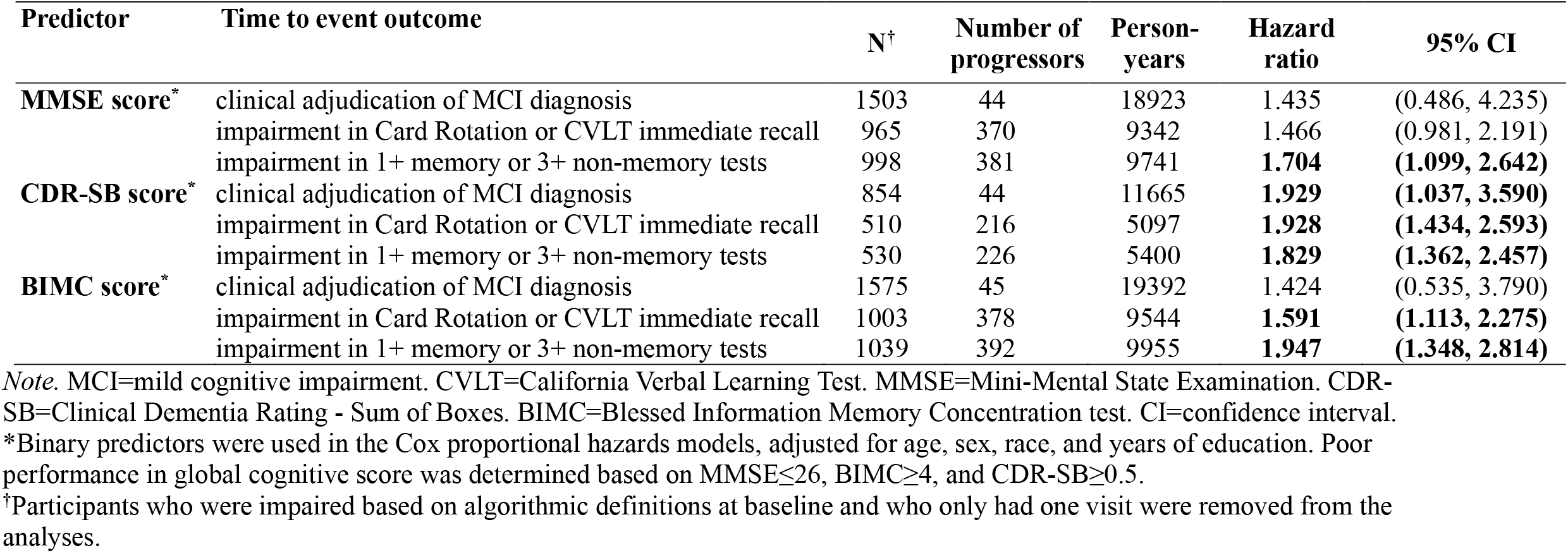
Baseline global cognitive scores as predictors of progression to algorithmically defined early cognitive impairment (ECI) during follow up.

| Predictor | Time to event outcome | N<sup>†</sup> | Number of progressors | Person-years | Hazard ratio | 95% CI |
| --- | --- | --- | --- | --- | --- | --- |
| **MMSE score** * | clinical adjudication of MCI diagnosis | 1503 | 44 | 18923 | 1.435 | (0.486, 4.235) |
|  | impairment in Card Rotation or CVLT immediate recall | 965 | 370 | 9342 | 1.466 | (0.981, 2.191) |
|  | impairment in 1+ memory or 3+ non-memory tests | 998 | 381 | 9741 | **1.704** | **(1.099, 2.642)** |
| **CDR-SB score** * | clinical adjudication of MCI diagnosis | 854 | 44 | 11665 | **1.929** | **(1.037, 3.590)** |
|  | impairment in Card Rotation or CVLT immediate recall | 510 | 216 | 5097 | **1.928** | **(1.434, 2.593)** |
|  | impairment in 1+ memory or 3+ non-memory tests | 530 | 226 | 5400 | **1.829** | **(1.362, 2.457)** |
| **BIMC score** * | clinical adjudication of MCI diagnosis | 1575 | 45 | 19392 | 1.424 | (0.535, 3.790) |
|  | impairment in Card Rotation or CVLT immediate recall | 1003 | 378 | 9544 | **1.591** | **(1.113, 2.275)** |
|  | impairment in 1+ memory or 3+ non-memory tests | 1039 | 392 | 9955 | **1.947** | **(1.348, 2.814)** |

*Note.* MCI=mild cognitive impairment. CVLT=California Verbal Learning Test. MMSE=Mini-Mental State Examination. CDR-SB=Clinical Dementia Rating - Sum of Boxes. BIMC=Blessed Information Memory Concentration test. CI=confidence interval.
\*Binary predictors were used in the Cox proportional hazards models, adjusted for age, sex, race, and years of education. Poor performance in global cognitive score was determined based on MMSE ≤ 26, BIMC ≥ 4, and CDR-SB ≥ 0.5.
<sup>†</sup>Participants who were impaired based on algorithmic definitions at baseline and who only had one visit were removed from the analyses.

Statistical tests were two-tailed and the significance level α was set as 0.05. All analyses were conducted using Stata version 16.1 (StataCorp, College Station, TX).

## Results

Among 1,851 participants in analyses, the mean age at baseline was 65.2 (SD=11.8) years, about half of participants were female (n=917, 49.5%), and 73.8% (n=1,366) were white **(Supplementary Table 1)**. The average education level was 16.8 (SD=2.7) years. At baseline with available cognitive data, over one third of the participants (n=628, 34.0%) were classified as having ECI based on poor performance in Card Rotations or CVLT immediate recall. One third of the participants (n=597, 32.3%) had ECI based on poor performance in memory or nonmemory tests.

### Concurrent criterion validation

**Table 1** summarizes evidence for concurrent criterion validity for both algorithmic classifications with respect to consensus diagnoses of MCI/dementia and global cognitive and functional scores. Compared with the consensus diagnoses, the AUC for the ECI algorithm based on poor performance in memory or nonmemory tests had a higher AUC (AUC = 0.703 vs AUC = 0.631). The two algorithms had comparable specificity (0.74 and 0.75) but the ECI algorithm based on memory or nonmemory tests had the higher sensitivity (0.65 vs. 0.52). Regarding evidence of concurrent criterion validity with MMSE, CDR-SB, and BIMC scores, the two algorithms had comparable AUCs, sensitivity, and specificity (**Table 1**).

### Predictive criterion validation

Among 1,851 participants, n=21 were diagnosed with MCI/dementia at baseline or prior BLSA visits. Among 1,538 participants with at least 2 visits with cognitive data and without MCI/dementia at baseline, 43 participants progressed to MCI/dementia. The average follow-up years between baseline and a consensus diagnosis of MCI/dementia was 12.3 (SD=6.9) years. **Table 2** summarizes the number of cases with consensus diagnoses over follow up and hazard ratios (HRs) for progression to MCI/dementia based on each algorithm. The algorithm based on impaired Card Rotations or CVLT immediate recall outperformed the other in terms of the ability to predict future progression to MCI/dementia. Participants with ECI based on this algorithm at baseline had over triple the risk of developing MCI/dementia (HR=3.53, 95% CI: 1.59-7.84) compared to those without ECI. The algorithm based on poor performance in memory or nonmemory tests also significantly predicted future progression to MCI/dementia (HR=2.24, 95% CI: 1.11-4.51).

### Global cognitive scores with future algorithmic determined ECI

We additionally examined the relationship between baseline MMSE, BIMC, and CDR-SB scores and hazard of early algorithmic diagnoses using Cox proportional hazards models **(Table 3)**. The algorithm based on poor performance in memory or nonmemory tended to have higher HRs of being predicted by MMSE or BIMC score. Comparable HRs were observed for the two algorithms when they were predicted by CDR-SB score.

## Discussion

This study contrasted two psychometrically defined algorithms to classify older adults with ECI in the BLSA. Results suggest that these ECI algorithms yielded comparably moderate concurrent criterion validity with consensus diagnoses of MCI/dementia and global cognitive and functional impairment. However, the algorithm based on poor performance in visuospatial ability (Card Rotations) or immediate memory (CVLT immediate recall) had a stronger relationship with future progression to MCI/dementia among the algorithms we evaluated. This pattern of findings indicates that impairment in visuospatial ability or memory may be capable of detecting early cognitive changes in the preclinical phase among cognitively normal individuals.

Our findings are consistent with previous research demonstrating that tests of visuospatial and memory function are among the earliest to show decline prior to onset of Alzheimer’s type dementia ^9,21^. Our results suggest that older adults with poor performance in Card Rotations or CVLT immediate recall had over triple the risk of progressing to MCI or dementia. Previous studies found that multiple domain amnestic MCI, defined as impairment in memory and at least one other domain (i.e., executive function, processing speed, language), significantly predicted incident dementia using data from the Framingham Heart Study ^5^. The algorithm based on one test in visuospatial ability and one test in episodic memory may provide novel and simplified neuropsychological criteria to identify ECI. Deficits in visuospatial functioning also have been associated with probable Lewy Body dementia ^30,31^. Studies also suggested the diagnostic and prognostic potential of visuospatial tasks in AD and non-AD dementias ^32–34^. The underlying mechanisms that may lead to early impairment in visuospatial ability as an indicator of ECI are related to the precuneus and other parietal regions that support spatial navigation ^35,36^. The precuneus is also one of the earliest brain regions to show β amyloid accumulation in preclinical AD ^37,38^. Our study extends previous research by demonstrating the predictive criterion validity of this algorithm with clinical diagnosis of MCI/dementia and highlights the importance of incorporating visuospatial ability into identification of ECI. Our findings may suggest a novel method to detect and diagnose cognitive impairment at an earlier stage. Future studies should investigate whether this algorithm is capable of identifying ECI in other older populations.

Great efforts have been made to define different neuropsychological criteria for MCI diagnoses and to derive a common classification algorithm for identification of MCI across several cohort studies ^4,6,8,39^. These efforts range from a single impaired memory score towards one or two tests in multiple cognitive domains such as memory, language, and speed/executive function ^6^, which enables identifications of MCI subtypes ^5,6^. The latter approach is consistent with DSM-5 criteria for mild and major neurocognitive disorder, which specifies domains of learning and memory, higher-level executive abilities, language, visuospatial function, and social cognition ^40^. Although, strictly speaking, our approach is not identical to the Jak/Bondi criteria which require at least two impaired scores (1 SD below the means) within a cognitive domain,^4,6^ we took both memory and non-memory tests into account. Future research is needed to validate these algorithms for early identification of cognitive impairment in other large cohort studies.

The algorithmic approach we describe may be useful in future clinical trials or observational studies as a validated tool to differentiate cognitively normal older adults who may be in the preclinical stages of AD, which may be an alternative to time-consuming adjudication. As these algorithms were derived based on age-, sex-, race-, and education-specific cut-points, this approach may be utilized in other cohort studies of cognitively intact older adults with diverse characteristics. Although the diagnosis of MCI in clinical settings also relies on other factors such as subject complaints and proxy reports, this study provides evidence for further investigations on application of algorithmic approaches as supplementary information in the clinical decision-making process.

Strengths of this study include large sample size, long follow-up period, and a large battery of neuropsychological tests. This study has several limitations. First, the generalizability of our findings to other cohorts needs to be considered in light of heterogeneity in cognitive batteries across studies. Validations of these algorithms using data from other cohort studies are needed. Second, the majority of BLSA participants were white. Although the algorithmic classification is race-adjusted, this approach should be validated in larger, more representative samples of diverse racial groups – especially given that sensitivity and specificity of algorithms varies across racial and other demographic groups ^41^. Third, we used an age-, sex-, race-, and education-adjusted cutoff of 1 SD to define poor performance on each test. Refinement of this cutoff is a viable future research area. Fourth, the algorithmic classification was determined based on age-specific means at any single time point. Although this approach may enhance clinical utility for providers with single office visit assessments, there is a possibility that an individual who was classified as impaired within their current age group may move to the unimpaired group in a subsequent visit, limiting the application of the algorithm to longitudinal studies aimed to examine changes in cognitive status. A further limitation is that while we are interested in a measure sensitive to cognitive changes in the preclinical phase of AD, our outcome was all-cause dementia as adjudicated by a clinical consensus committee, which is a broader classification than AD as a primary etiology. We did not explore that outcome because of the confluence of small case numbers coupled with misclassification errors in diagnoses of living participants. ^42^

In conclusion, this study compared two classification algorithms to detect early cognitive impairment among cognitively normal adults aged 50 years and older enrolled in the BLSA study. The algorithm based on impairment in visuospatial ability or immediate recall had a stronger relationship with future progression to consensus diagnoses of MCI or dementia. These algorithmic approaches may be further utilized to detect early cognitive changes in the preclinical phase before progression to symptomatic phase of dementia. Future studies incorporating motor function impairment into the algorithms may further enhance the ability to capture preclinical changes across the spectrum of various types of dementias. Additional research is needed to relate the algorithmic approaches to AD biomarkers and apolipoprotein E (APOE) genotype.

## Data Availability

All data produced in the present study are available upon reasonable request to the authors

## Acknowledgments

This work was supported by the National Institutes of Health (NIH) grant R01AG061786. Y.C. and J.A.S. were supported by R01AG061786 and U01AG057545. A.G. was supported by the NIH K01AG050699. This study was also supported in part by the Intramural Research Program, National Institute on Aging, NIH. N.M.A., M.K-T., L.F., E.M.S., S.M.R. were all supported by the Intramural Research Program of the National Institute on Aging, NIH.

