## Supplementary Table 1 for "Application and validation of an algorithmic classification of early impairment in cognitive performance"

**Supplementary Table 1. Sample characteristics at baseline**

| Characteristics | All |
| --- | --- |
|  | N=1,851 |
| Age (years) | 65.2 ± 11.8 |
| Female | 917 (49.5) |
| White | 1366 (73.8) |
| Education years | 16.8 ± 2.7 |
| Adjudicated MCI/dementia | 21 (1.1) |
| ECI algorithms |  |
| Impairment in Card Rotation or CVLT immediate recall | 628 (34.0) |
| Missingness | 5 |
| Impairment in 1+ memory or 3+ non-memory tests | 597 (32.3) |
| Impairment in cognitive tests |  |
| CVLT immediate recall | 347 (19.1) |
| Missingness | 36 |
| CVLT long-delay free recall | 397 (22.0) |
| Missingness | 42 |
| Card Rotations test | 382 (21.4) |
| Missingness | 69 |
| BVRT | 309 (16.8) |
| Missingness | 12 |
| Digit backward | 310 (17.0) |
| Missingness | 32 |
| Digit forward | 354 (19.5) |
| Missingness | 34 |
| Boston Naming Test | 191 (19.4) |
| Missingness | 868 |
| Similarities | 296 (19.8) |
| Missingness | 359 |
| Average cognitive tests scores |  |
| CVLT immediate recall | 51.6 ± 11.7 |
| CVLT long-delay free recall | 10.6 ± 3.5 |
| Card Rotations test | 80.7 ± 38.4 |
| BVRT (reverse coded) | 29.7 ± 4.6 |
| Digit backward | 7.2 ± 2.3 |
| Digit forward | 8.3 ± 2.3 |
| Boston Naming Test | 53.3 ± 6.5 |
| Similarities | 20.9 ± 3.9 |

*Note.* MCI=mild cognitive impairment. ECI=early cognitive impairment. CVLT=California Verbal Learning Test. BVRT=Benton Visual Retention Test.
